# Remote and semi-automated methods to conduct a randomized clinical trial

**DOI:** 10.1101/2022.11.08.22281952

**Authors:** Teresa Cafaro, Patrick J. LaRiccia, Brigid Bandomer, Helen Goldstein, Tracy L. Brobyn, Krystal Hunter, Satyajeet Roy, Kevin Q. Ng, Ludmil V. Mitrev, Alan Tsai, Denise Thwing, Mary Ann Maag, Myung K. Chung, Noud van Helmond

**Author notes:** **Corresponding Author:** Teresa Cafaro, One Cooper Plaza, Camden, NJ 08103, USA.

## Abstract

**Background:** Designing and conducting randomized controlled trials is not possible for some institutions and researchers due to their associated costs and personnel requirements. Advances in telehealth for patient care and clinical trial methods adaptations were hastened due to the coronavirus 2019 pandemic. We conducted subject recruitment, screening, informed consent, study product distribution, and data collection remotely using Research Electronic Data Capture (REDCap) and other readily available applications for a prospective randomized clinical trial on vitamin D3 supplementation to prevent influenza-like illness in healthcare workers.

**Objectives:** To describe how to efficiently and cost-effectively conduct a randomized clinical trial using remote and semi-automated methods.

**Methods:** A previously described prospective, controlled, trial in healthcare workers at a tertiary university hospital that used Zelen’s design to facilitate enrollment is used as a model. A random group of healthcare workers were invited to participate in the study through email. Following an automated process, interested individuals could schedule a HIPAA compliant, video-facilitated consent interview. Following e-consenting, participants received vitamin D3 supplementation bottles by mail and electronic surveys via email. Adherence to the study intervention was monitored through survey data review and vitamin D3 safety was monitored via real-time automated warnings to study staff if any symptoms, health changes, hospitalizations, or pregnancies were reported on monthly surveys.

**Results:** A small study staff of 10 team members were able to screen 406 subjects and enroll 299 subjects over a 3-month period using completely remote methods. Adherence to vitamin D3 supplementation was high (87%) over 9 months. Survey data completeness was 98.5% over 9 months. Participants and study staff scored the system usability 93.8% and 90%, respectively. The automated and remote methods allowed the study maintenance period to be managed by a small study team staff of 2 members while safety monitoring was conducted by 3-4 team members.

**Conclusions:** The remote and automated methods developed to conduct a randomized controlled trial produced efficient subject recruitment with excellent study product adherence and data completeness. These methods can significantly minimize costs and time without sacrificing safety or quality, thereby offering greater equity in scientific research. We share our XML file for researchers to use as a template for learning purposes or designing their own clinical trials.

**Trial Registration:** NCT04596657

## INTRODUCTION

Designing and conducting randomized controlled trials, the gold standard of research,^1^ appears to be out of reach for many institutions and independent researchers due to the exorbitant costs, personnel requirements, and extensive time it takes to complete such endeavors.^2^ Enrollment in prospective clinical trials was significantly hampered due to pandemic restrictions on non-urgent contact with patients and reallocation of resources to COVID-19 (coronavirus disease 2019) care.^3^ Technological advances in patient care via telehealth visits emerged rapidly and were promptly adopted by healthcare systems and reimbursement organizations.^4,5^ As with clinical care, there was an urgent need to leverage technological advances to conduct prospective clinical trials remotely.^3,6^ Research Electronic Data Capture (REDCap) was utilized by several groups to conduct their trials both prior to^7-10^ and during the pandemic.^11-13^ Our trial combined REDCap and other electronic applications to reduce many of the logistical requirements involved in executing a clinical trial.

During the coronavirus pandemic, we conducted a prospective randomized trial in healthcare workers on the protective effect of vitamin D supplementation on influenza-like illness.^14^ We herein describe electronic communication, study product shipping, and data capture methods to conduct a randomized controlled trial remotely using readily available software packages and platforms. We propose that these pragmatic methods can be used to efficiently and cost-effectively conduct clinical trials without sacrificing safety or data quality.

## METHODS

### Study Overview

A randomized clinical trial on the protective effect of vitamin D supplementation in healthcare workers on developing influenza-like illness, including COVID-19, was conducted in a large tertiary academic hospital. The institutional review board approved the study (IRB #20-455) which followed a Zelen’s design wherein only participants randomized to the intervention group were approached for participation. Zelen’s design is ethical and useful within the context of trials of prevention and screening interventions.^15,16^ The trial flow is presented in the CONSORT flowchart – Figure 1. All participants who received vitamin D3 provided electronic informed consent. All study information and trial logistics were managed using the Research Electronic Data Capture (REDCap) system version 10.8.2 developed by Vanderbilt University.^17,18^ The REDCap Automated Survey Invitations (ASI), branching logic, piping feature and alerts were heavily utilized for automation along with integrated applications for texting and appointment scheduling. All study contacts with subjects, including recruitment, consent, data collection and safety monitoring were virtual, with the exception of clinically indicated outpatient visits with the study safety monitor (investigator SR). An overview of all remote procedures is provided in Figures 2a-c.

**Figure 1.**
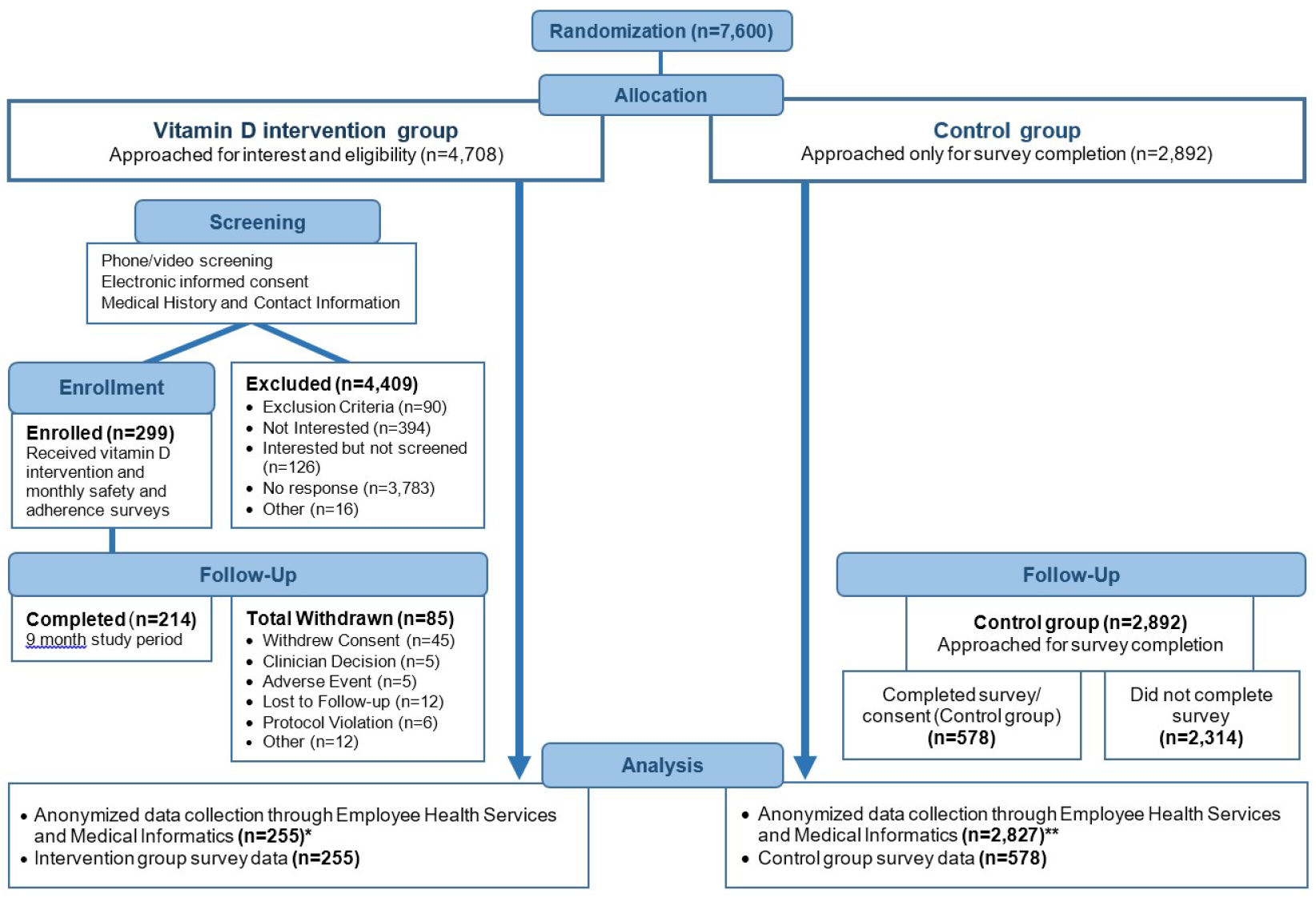
CONSORT trial flowchart. *Out of 299 intervention subjects, 44 subjects were excluded from analysis because they were not on 5,000 IU vitamin D3 per day for at least 60 days, a timeframe that is considered to be protective. ^29^ The resulting 255 subjects included subjects who completed the 9-month study period and early withdrawals. ** Employee Health was unable to locate 27 control subject records for unknown reasons. A possibility for missing records could be due to name changes over the course of the study. Another 38 control subjects were found to have been terminated from employment prior to the study observation period and were excluded from analyses (n = 2,892 - 27 - 38 = 2,827).

**Figure 2a.**
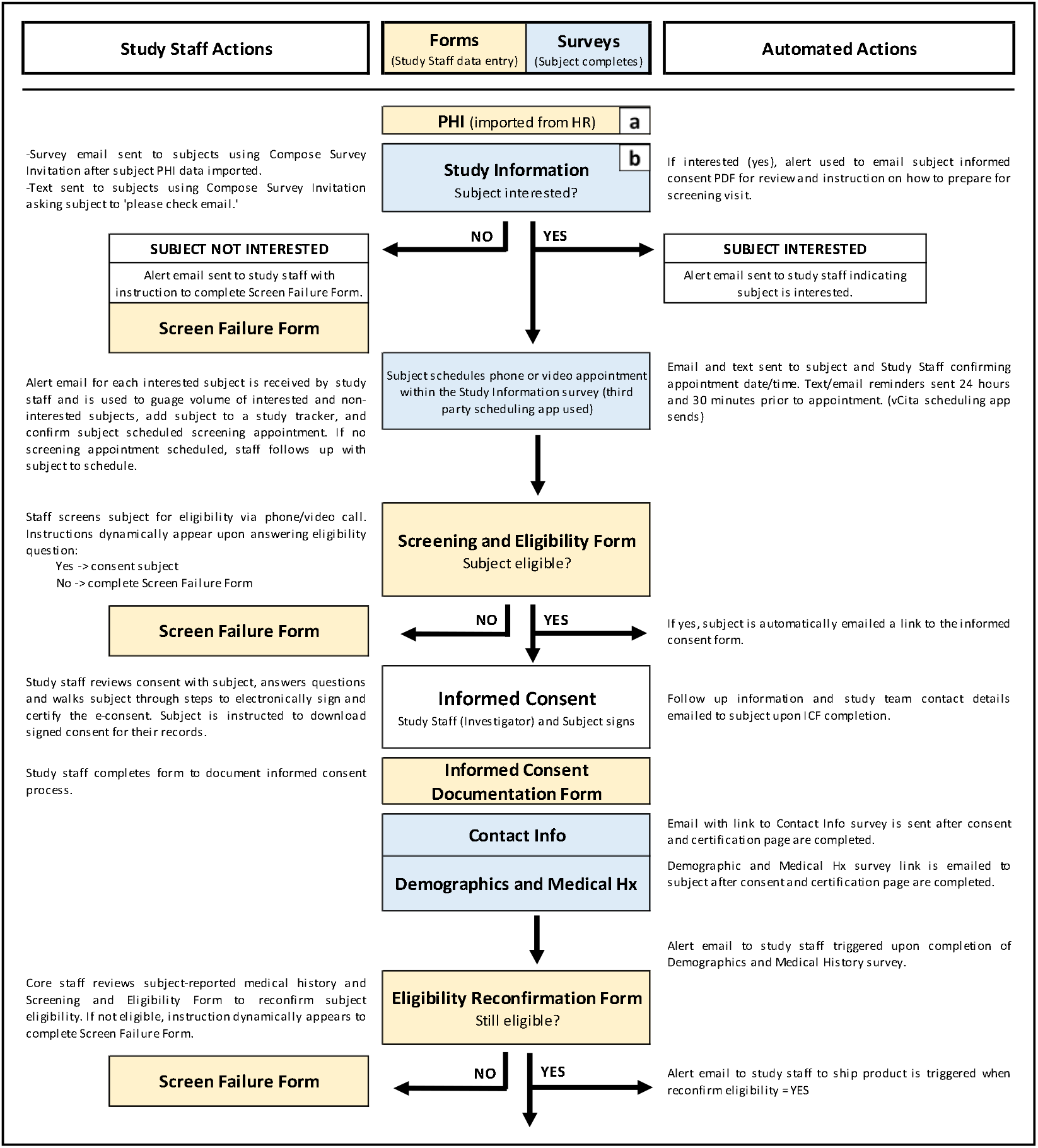
Surveys and forms flowchart - screening and enrollment period. REDCap instruments are used for data collection and can be created as **forms** to be completed by study staff (yellow) or **surveys** to be completed by subjects (blue). a) With Institutional Review Board (IRB) approval and Human Resources (HR) support, healthcare employees were randomized prior to sending an invitation to participate. Employee names and emails provided by Human Resources were uploaded to the PHI form (Protected Health Information). b) The Study Information survey was created to invite employees to participate; allow potential subjects to schedule a phone or video screening appointment if interested; provide a summary of study details with text and video; and offer the full informed consent should potential subjects wish to review it. See Figure 3a for more detail on the Study Information survey. Before sending the Study Information survey, emails were sent to all employees informing them of the upcoming study.

**Figure 2b.**
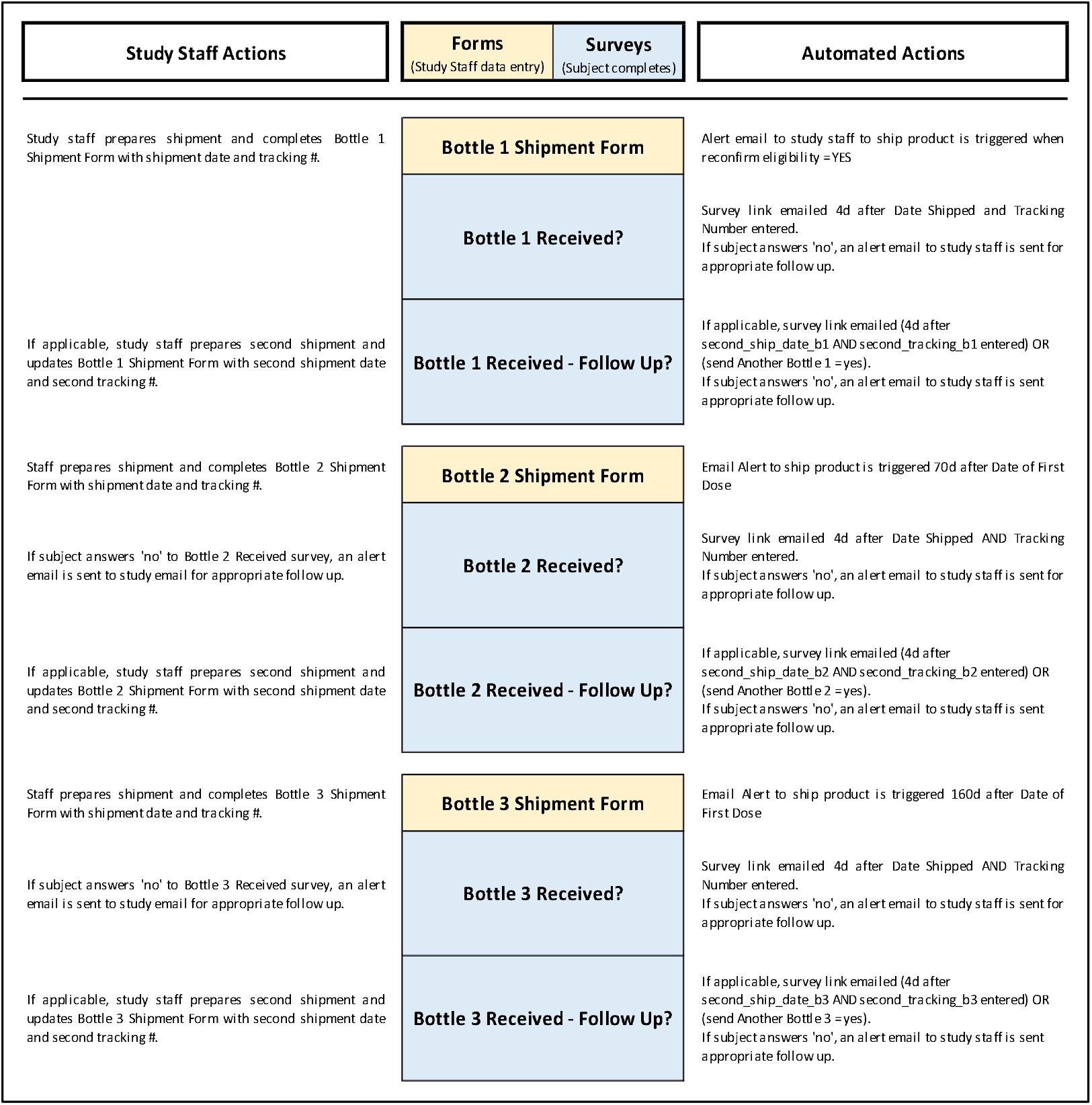
Surveys and forms flowchart – study product shipment and tracking. Study product was supplied to subjects in 3 separate bottles containing 90 gelcaps each (Bottle 1, Bottle 2, Bottle 3) with the aim of easing pill counts for the subjects at month 3, month 6 and month 9 timepoints. The Bottle Shipment forms were created to capture shipment dates, carrier tracking numbers; to remind the staff member to include all items in the initial shipment (study product, pill organizer and study packet letter); and to include special delivery instructions for the carrier, if any. Each Bottle Shipment form (see Panel d in Figure 6a and Figure 6b) has piping in place (blue text) which displays subject shipping details on the form, thereby improving efficiency by removing the need to navigate to another form for the subject address. After email alert is received, study staff completes the Bottle Shipment Form and ships product. Four days after the shipment date, the Bottle Received? survey link is automatically sent to the subject. If subject indicates the bottle was not received, an email alert is sent to study staff prompting follow up and completion of the second section of the Bottle Shipment form which will set off the automated sequence just described, but for the second replacement bottle being shipped (i.e., alert to study staff to ship product; 2^nd^ bottle is shipped and 2^nd^ date of shipment is entered on the Bottle Shipment Form; email with link to Bottle 1 Received - Follow Up? survey sent to subject; subject indicates if bottle received; etc.).

**Figure 2c.**
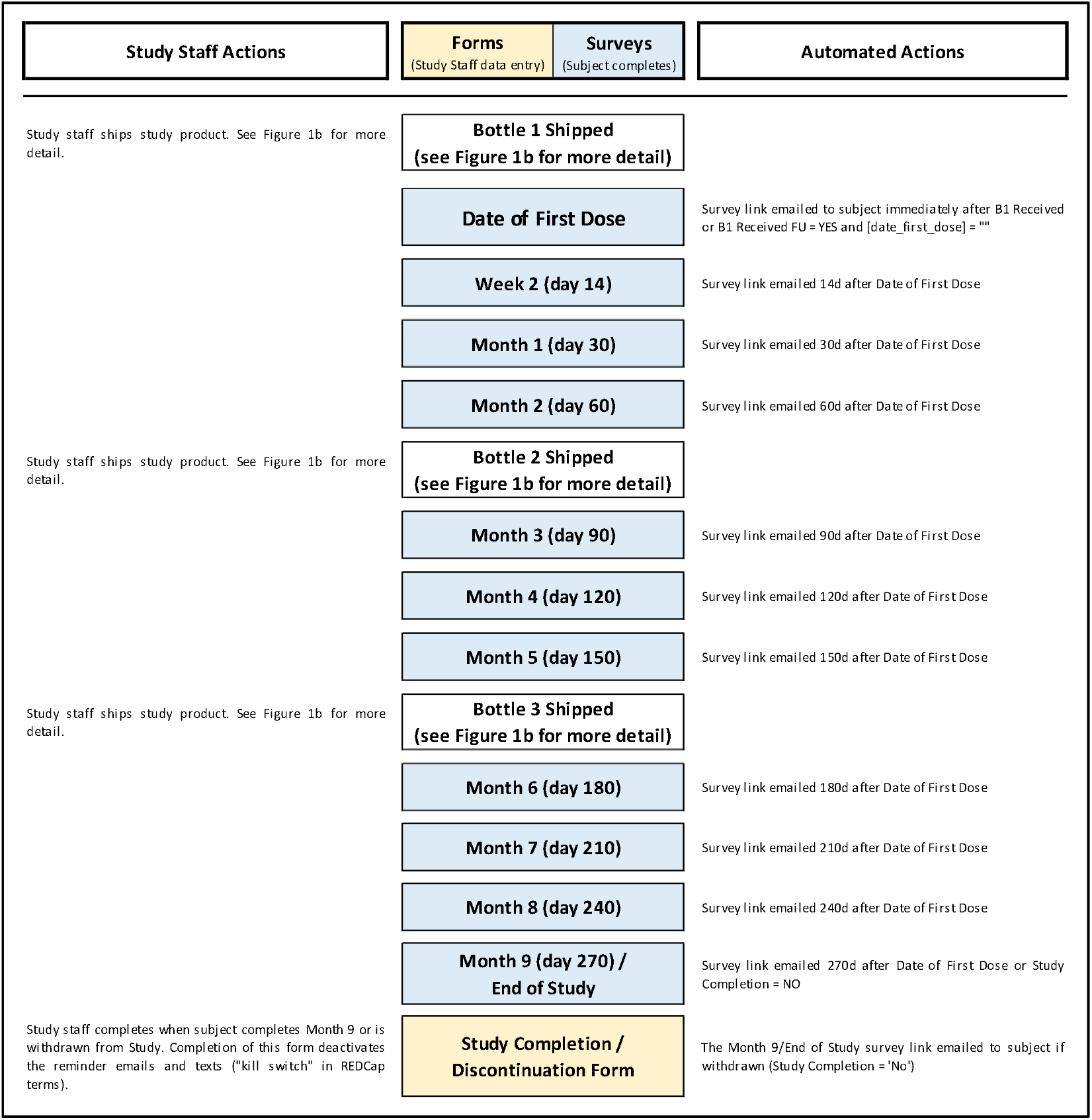
Surveys and forms flowchart – active study period. This flowchart illustrates the monthly surveys and the schedule by which the survey links are sent to subjects. REDCap Automated Survey Invitations (ASI) are created for each survey and use conditional logic to send surveys at the specified timeframe after the Date of First Dose. In addition, a “kill switch” was incorporated in the conditional logic for each ASI: if the Study Completion / Discontinuation Form is completed the survey will not be sent. Example of conditional logic for Month 3 survey: datediff ([enrollment_arm_1] [date_first_dose], ‘today’,’d’, true) = 90 and [month_9_arm_1] [complete_study_eos] <> ‘0’. For more complex logic used for study product shipment and tracking, see Figure 2b. In addition, the XML file containing all conditional logic is shared in the Data Availability section.

### Randomization

Seventy-six hundred employees (subjects) were randomly assigned study numbers and divided into two groups: 1) the intervention group (vitamin D supplementation); and 2) the control group. Under this study design randomization was performed prior to recruitment and screening. Randomization was completed using the randomization function in Microsoft Excel. The intervention group was approached for study participation. The control group did not receive placebo and was approached for voluntary survey completion toward the end of the 9-month intervention study period to compare the two groups for demographic and clinical characteristics.

### Recruitment

#### Intervention and control groups

As recruitment by email can be challenging, we opted to inform subjects of the study prior to sending the REDCap-generated email containing the study invitation link. First, a bulletin summarizing the study and informing employees of potential contact was posted on our institutional intranet. Subjects were made aware of the posting via a weekly email announcement from institutional leadership.

Second, the principal investigator emailed subjects informing them the forthcoming study invitation email was not fraudulent and the study invitation link was safe to open. The study was led by health care system staff; and participation was voluntary.

### Screening & Electronic Informed consent

#### Intervention group

The intervention group subjects were emailed a link to the study information survey through REDCap. This survey contained summarized study information, a brief informational video about the study, and a full PDF of the informed consent. Subjects were asked to indicate their potential interest in the study (Figure 3a). If interested, instructions dynamically appeared for the subjects to electronically schedule their screening and consent appointment (Figure 3b). We used a third-party Health Insurance Portability and Accountability Act (HIPAA)-compliant appointment scheduler (vCita, Bellevue, WA, USA) developed for telemedicine appointments. The scheduling application was integrated within REDCap using an application programming interface (API). Both telephone and video call (Zoom Video Communications, San Jose, CA, USA) appointments were offered. The scheduling application allowed for study team members to adjust their availability for appointment assignments. After a potential subject booked a screening appointment, both the subject and study team member were sent an automated calendar invitation for the appointment with the relevant contact information or video call link. In addition, the study team member was informed of the scheduled appointment via text in real time. Through the scheduling application, the potential subject was automatically sent a PDF copy of the informed consent form (without signature section) for their review before the screening appointment as well as instructions on how to prepare for the screening appointment. Instructions for the appointment included reviewing the informed consent form, having information on any medications or supplements they were taking available, and being seated at a computer/tablet/smartphone with access to internet and email. The scheduling application was set to send reminder emails 24 hours and 30 minutes prior to each appointment.

**Figure 3a.**
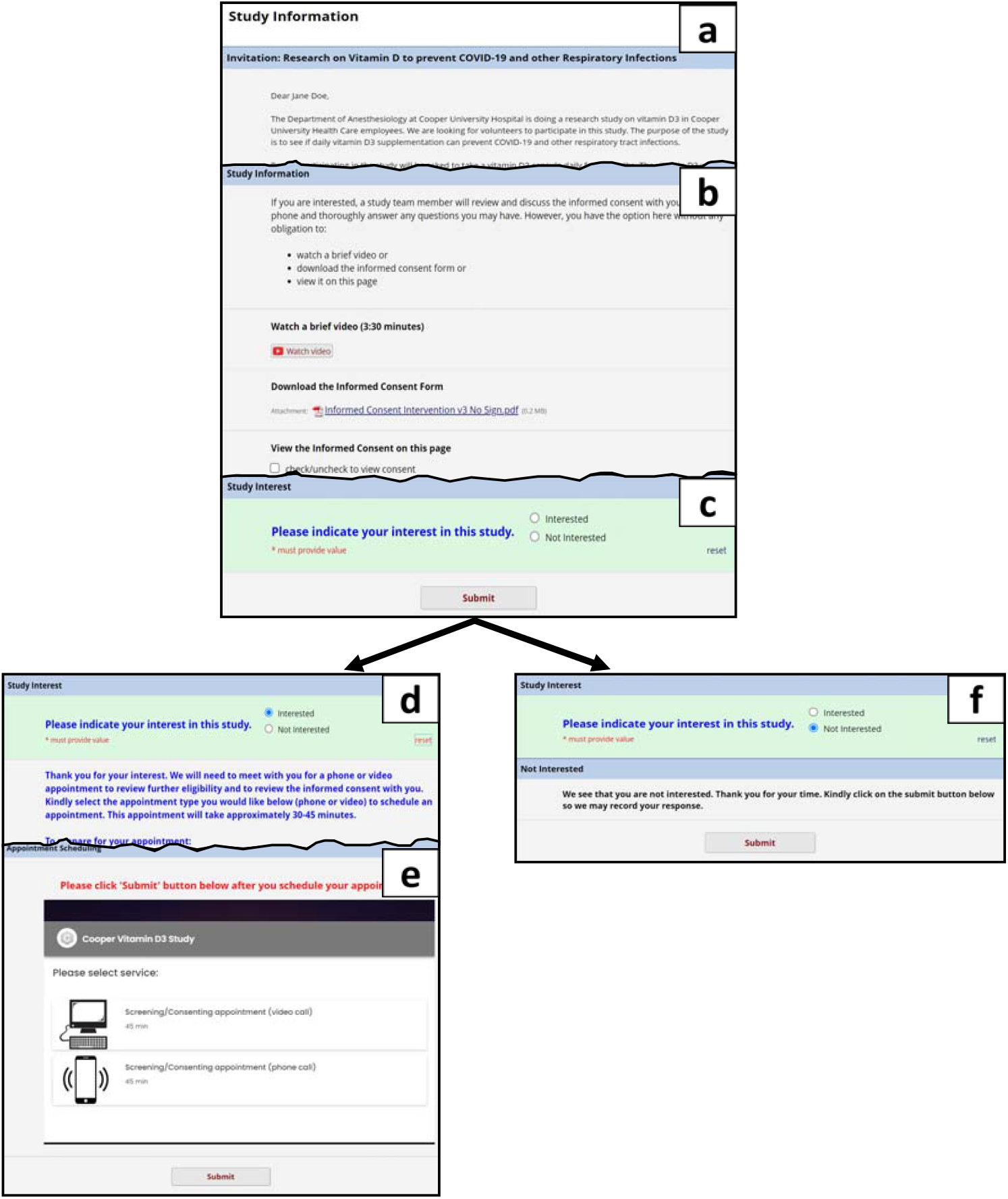
Study information survey. Panel a: Invitation with brief explanation. Panel b: Study information with option to watch brief video, download consent, or view consent inline. Panel c: Study interest question which, depending on the answer, will dynamically reveal next steps and relevant instructions. Panel d: If interested, instructions for screening appointment preparation and electronic appointment scheduling dynamically appear. Panel e: Electronic scheduler integrated with REDCap using API. See Figure 3b for more detail on electronic scheduler. Panel f: If not interested, a thank you to the subject with a request to submit survey dynamically appears.

**Figure 3b.**
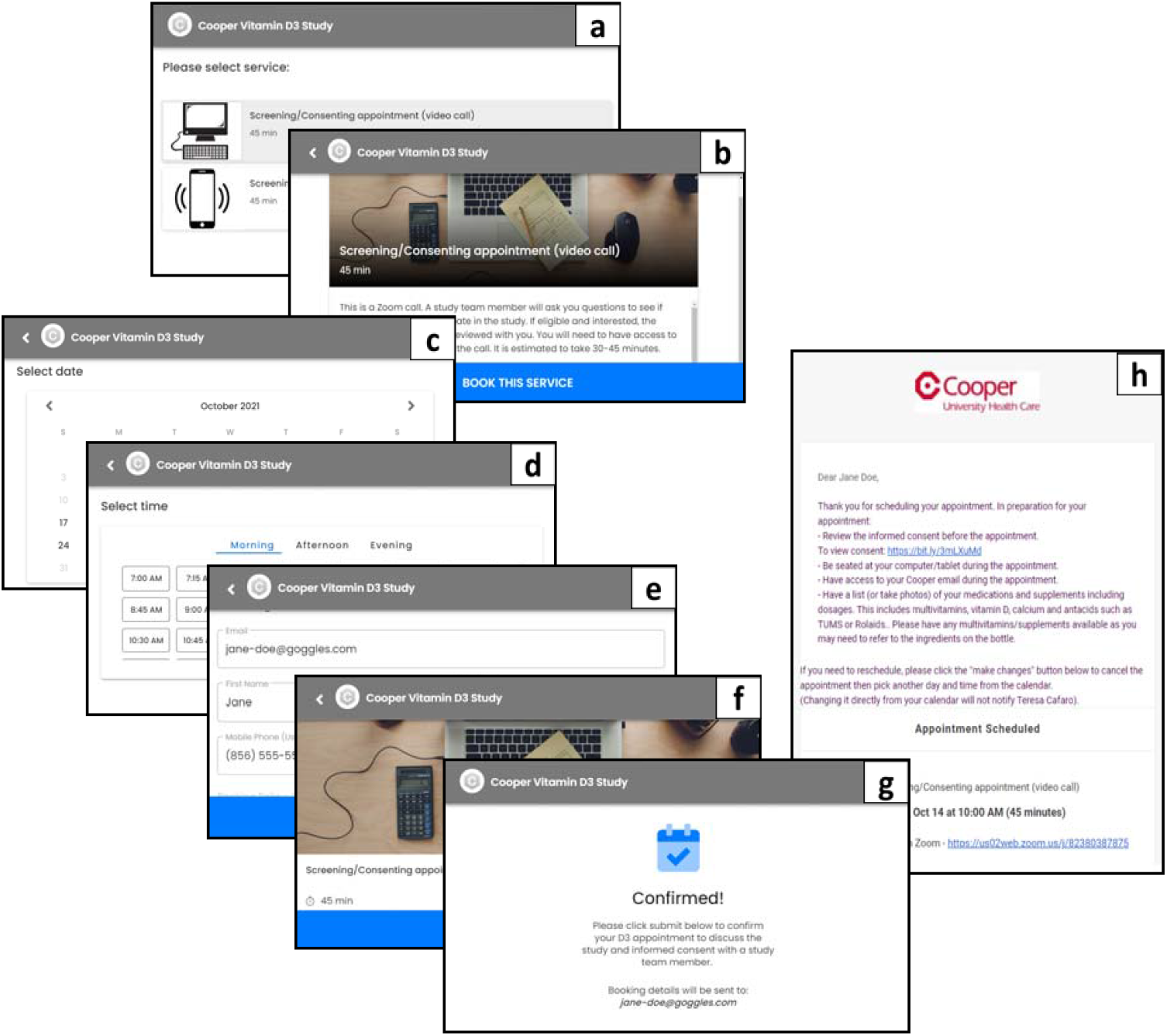
Study information survey - Electronic appointment scheduling. The Study Information Survey (Figure 3a) included an electronic appointment scheduler for subjects to schedule their screening and consent appointment at an available time slot. We illustrate here an overview of the scheduling process for the subject. A third party HIPAA-compliant appointment scheduler (vCita, Bellevue, WA, USA) which was developed for telemedicine appointments was used. The scheduling application was integrated within REDCap using API (Application Programming Interface). Both telephone and video call (Zoom Video Communications, San Jose, CA, USA) appointments were offered (Panels a, b). The scheduling application allowed for study team members to adjust their availability for appointment assignments (not shown). After a potential subject booked a screening appointment (Panels c, d, e, f, g), both the potential subject and study team member were sent an automated calendar invite for the appointment with the relevant contact information or video call link (Panel h). The study team member was also informed of scheduled appointment via text. In addition the potential subject was automatically sent a PDF copy of the informed consent form (without signature section) for their review before the screening appointment as well as instructions on how to prepare for the screening appointment. Instructions for the appointment included reviewing the informed consent form, having information on any medications or supplements they were taking available, and being seated at a computer/tablet/smartphone with access to internet and email. The appointment scheduler was set to send reminder emails 24 hours and 30 minutes prior to each appointment.

During the scheduled appointment, a study team member described the study in detail. If a subject confirmed interest, eligibility was determined through a screening and eligibility script which focused on exclusion criteria related to medical history and medication use. During the call, the study team member completed a screening and eligibility form in REDCap (Figure 4). Depending on the information entered, relevant instructions dynamically appeared for the team member (e.g., if eligibility = no, subject is a screen failure; proceed to the Screen Failure Form to record pertinent information). Eligible subjects who wished to proceed with the consent interview (e.g., eligibility = yes) were automatically sent an email with a link to the subject-specific informed consent. Subjects were asked to open the informed consent form from their email on a computer, tablet, or smartphone. The informed consent form was reviewed in detail, and teach-back questions were used to confirm the subject’s understanding of the study. After reviewing the consent and answering any questions, the study investigator instructed the subject on the steps needed to electronically sign and date-timestamp the informed consent form [Supplement 1]. The electronic consent was adapted from Combined Consent & HIPAA Authorization Template as posted in the REDCap library.^19^ We used the electronic consenting framework in REDCap (Figure 5) which has an Auto-Archiver and e-Consent Framework wherein a PDF of the signed consent is automatically archived in REDCap’s file repository. The subjects’ name and consent version were automatically included on the footer of each page as extra documentation of the identity of the subject who is consenting. Subjects had two opportunities to download the signed informed consent form during the process. Alternatively, subjects were emailed a fully signed copy of the informed consent form through secure email.

**Figure 4.**
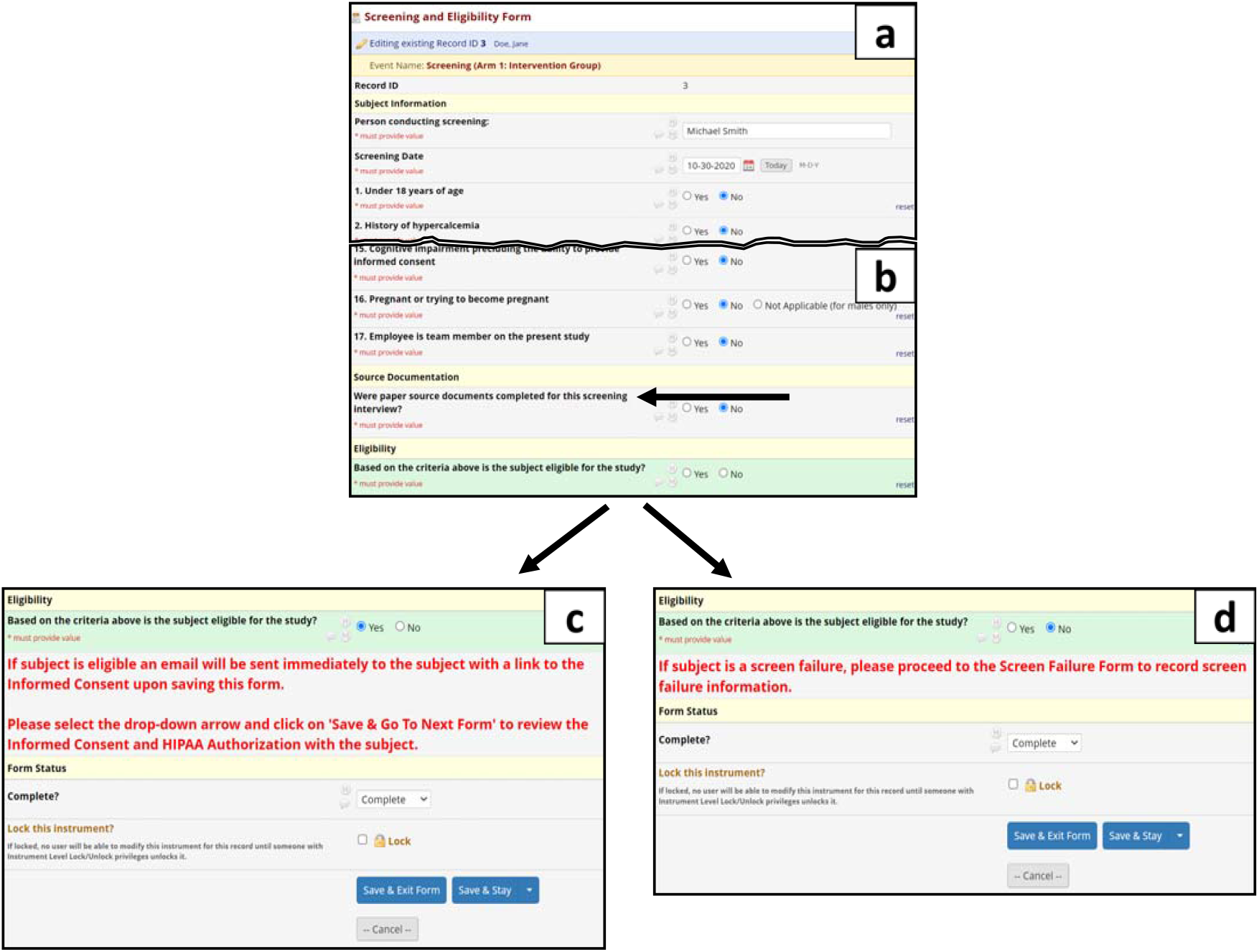
Screening and eligibility procedures. Panel a: Study team screened subjects with the aid of the Screening and Eligibility form. A Screening and Eligibility Phone Script was provided for use as source documentation or as a reference tool. Panel b: Team member chose direct data entry or paper source documentation. The eligibility question, however, must be answered “yes” at the time of screening to move onto the electronic consenting process. If source documents are used (=yes), instructions dynamically appear (not shown): “Please upload source documents into the REDCap File Repository. Your paper source documents will need to be sent to Cooper Anesthesiology Department for filing/storage at the end of the screening and enrollment period.” Panel c: Utilizing REDCap branching logic, specific instruction is provided to study team if subject is eligible (=yes) and wishes to review/sign the informed consent. Panel d: If subject is not eligible (=no) instruction to complete the screen failure form dynamically appears.

**Figure 5.**
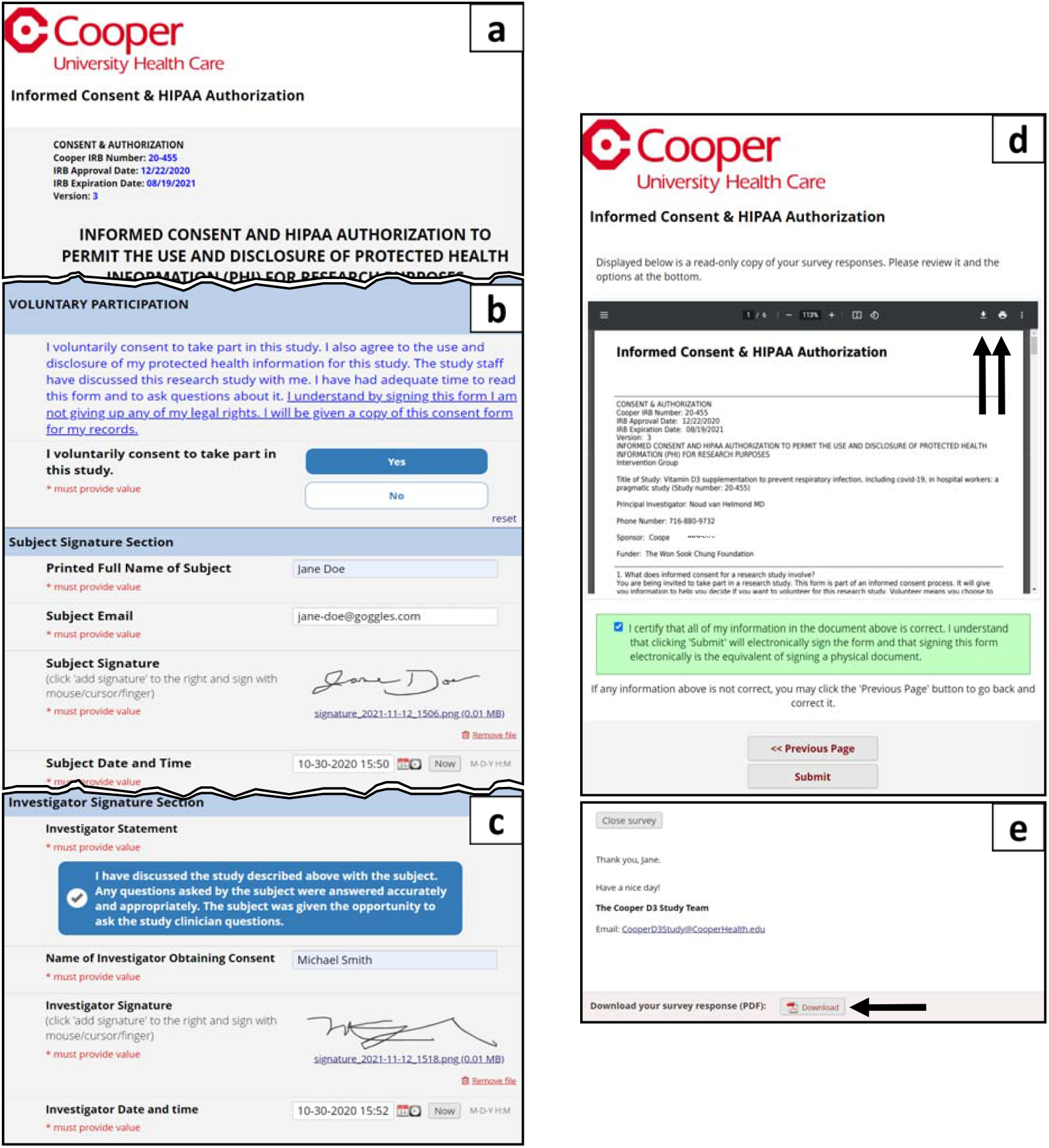
Electronic informed consent (e-consent). Panel a: Thee-Consent process was approved by our local IRB. The REDCap electronic consent framework (an option in survey settings) was utilized to consent and collect signatures from subjects and study staff remotely. The survey formatting was modified from the REDCap Shared Library. Panel b: The subject reads the voluntary consent statement; indicates if they voluntarily consent to participate; provides signature, email and date/time; and saves the document to allow for the investigator to sign and date/time the consent. Panel c: The investigator reviews and checks the investigator statement; enters name; provides signature and date/time; and saves entries. The subject refreshes the webpage, scrolls to bottom, confirms both subject and investigator signatures, and clicks ‘Next Page’ (not shown). Panel d: The consent framework includes a separate certification page. The subject is asked to review the inline signed consent; instructed on how to download or print the signed consent; and asked to certify that details are correct. Upon submitting the survey a PDF of the consent is automatically archived in the file repository (not shown). Panel e: After submission, subject had a second opportunity to download PDF of signed consent. Immediately following informed consent, subjects were emailed study team contact information, a link to the Demographics and Medical History form and a link to the subject Contact Information form. Supplement 1 is available detailing the steps the subject and investigator took to sign the informed consent.

#### Control group

Healthcare workers randomized to the control group were invited via email with a link to complete a survey. The control survey contained a summary of the study and an in-line scrollable informed consent form, which they were asked to read if they were interested in participating. An opportunity to ask questions about the study was offered by providing an option to call the 24/7 study phone number. If subjects decided to participate, they consented by answering a question about voluntary participation and providing their date- and time-stamped electronic signature. Subjects were asked to download the consent for their records.

### Enrollment

Signing the consent immediately triggered two surveys to be sent to subjects: 1) a contact information survey which asks subjects to confirm their name, preferred email, phone number, and home address for study product shipment; and 2) a demography and medical history survey which collected demographics, height, weight, history of low vitamin D level, if known, medical history and concomitant medications. Once the surveys were completed, an alert was sent prompting a study investigator to review the detailed medical history and to confirm that the participants still qualified and no answers conflicted with the information provided during screening. Once eligibility was reconfirmed, an email alert was emailed to a team member to ship the study product. The automated enrollment steps are detailed in Figures 6a-b.

**Figure 6a.**
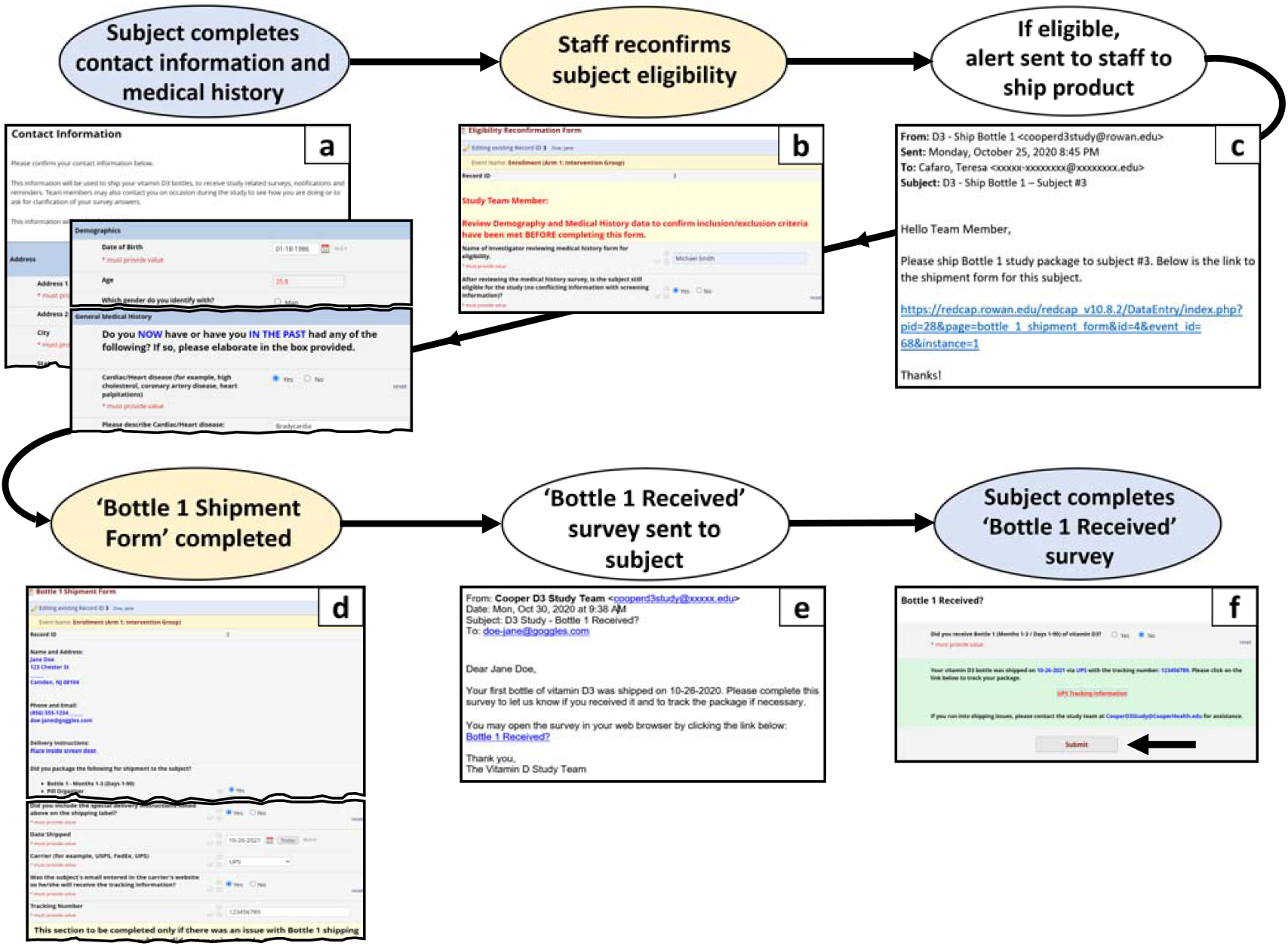
Automated enrollment steps. (Blue=survey; yellow=form; white=automated action) This schematic details the steps after consent is signed. After consent is signed and submitted, subject is auto-sent 2 emails with survey links using Automated Survey Invitations (ASI): Contact Information survey and Demographics and Medical History survey (Panel a). Once subject completes the surveys an alert email (not shown) is triggered to core study staff with instruction to review medical history and reconfirm subject eligibility (Panel b). If not eligible (=no), dynamic instructions appear to complete the Screen Failure form (not shown). If eligible (=yes), an alert email with a link to the Bottle 1 Shipment form is automatically sent to study staff notifying them to ship the first bottle of study product (Panel c) and complete the Bottle 1 Shipment form (Panel d). The Bottle 1 Shipment form has piping in place (blue text) which allows the staff member to see the subject shipping details on the form, thereby improving efficiency by eliminating the need to navigate to another form. Four days after product is shipped, an automated survey invitation (Panel e) is sent to the subject to request completion of the Bottle 1 Received survey (Panel f). If the package was not received (=no), the survey dynamically provides the shipment tracking link for the subject and subsequently an alert email is sent to study staff informing them of shipping issue.

**Figure 6b.**
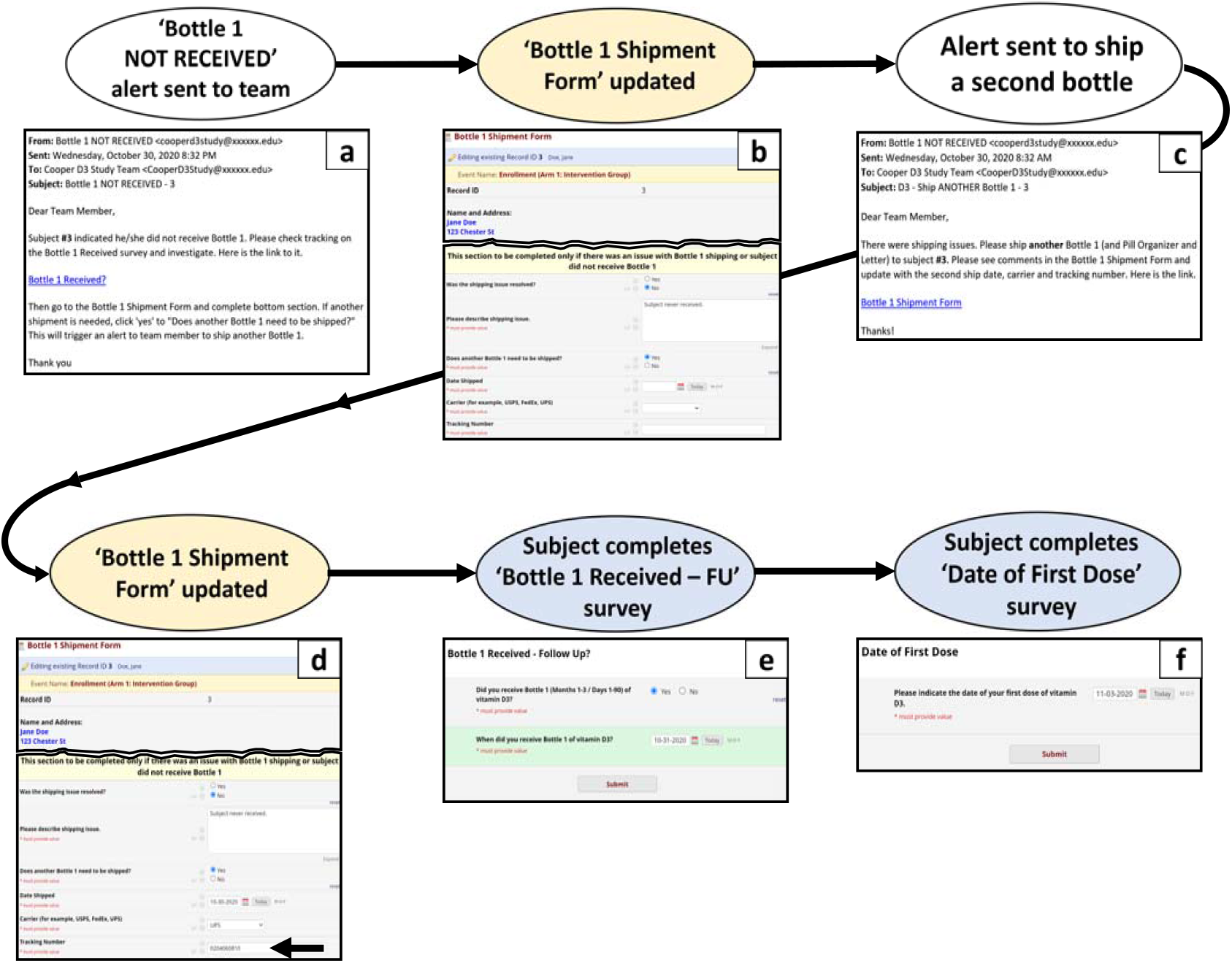
Automated enrollment steps. (Blue=survey; yellow=form; white=automated action) If bottle was not received, an alert email was sent to study staff informing them of a shipping issue and directing them to investigate (Panel a). After staff confirmed a second bottle must be shipped, they answered the first 3 questions of the last section of the Bottle Shipment form (Panel b). By answering ‘yes’ to ‘Does another Bottle 1 need to be shipped?’ an alert (Panel c) was automatically sent to designated study staff requesting they ship another bottle and complete the date shipped, carrier and tracking number fields on the Bottle 1 Shipment Form (Panel d). Four days after product was shipped, an automated survey invitation was sent to the subject requesting completion of the Bottle 1 Received – Follow Up survey (Panel e). If the package was received (=yes), a question asking for the date received dynamically appeared. Once submitted, the Date of First Dose survey was immediately triggered and sent to the subject. A study staff member monitored completion of Date of First Dose surveys dose and followed up with subjects to request completion. Thereafter, surveys were automatically sent as scheduled using the date of first dose as shown in Figure 2c.

#### Control group

Immediately after signing the consent, subjects proceeded to answer survey questions regarding demographics, medical history, use of vitamin D supplementation, vaccination history, and concomitant medication.

### Study product shipping

#### Intervention group only

The automated study product shipment details are depicted in Figures 2b and 6a-b. Study product was provided to subjects in three separate shipments over the 9-month study period. The first shipment was a study packet including 1) a 90-day supply of vitamin D3 (5,000 IU capsules daily); 2) a Study Packet Letter providing study information and instructions; and 3) a 7-day pill organizer to aid in adherence to study product. The study packet was shipped upon enrollment and two subsequent bottles were shipped at 3-month intervals. Shipment alerts were automatically sent to study staff based on the subjects’ study day threshold (i.e., day 0, day 90, day 180) and if their study status remained active.

When shipment notifications were received by study staff, subjects’ information was entered in the carrier’s electronic shipping form which created tracking numbers and address labels. Tracking numbers were entered into the REDCap shipping form along with date shipped and person shipping. Labels were applied to premade packages that were then delivered to the shipping company. All notifications and alerts for shipments or repeat shipments were through REDCap.

Four days after product shipped, an automated ‘Bottle Received?’ survey was emailed to subjects. A link to the shipment tracking number dynamically appeared to subjects if they answered ‘no’ on the survey. Answering ‘yes’ triggered the Date of First Dose survey link to be sent to subjects. Because the shipment tracking link was only visible on the survey if subjects answered ‘no’, the shipment tracking link was later inserted into the Bottle 2 and 3 shipment *forms* that staff completed and often referenced. The sample shipment tracking link utilized HTML coding:

<a target=“_blank” href=“https://www.ups.com/track?loc=null&tracknum=[month_2_arm_1][tracking_b2] &requester=WT/”>UPS Tracking Information</a>

### Survey data collection

#### Intervention group

At 2 weeks, 1 month, 2 months, 3 months, 4 months, 5 months, 6 months, 7 months, 8 months, and 9 months after date of first dose, an adherence and safety survey was sent as a link via email. ASIs were utilized to email subjects links to each survey at the study-defined timepoints (14 days, 30 days, 60 days, 90 days, etc).

Adherence to survey completion is necessary for robust data collection thus we employed methods to increase this adherence. If the survey was not completed within 24 hours, an automated reminder was sent daily for up to 3 days. If an individual survey was not completed after 3 reminders, a study team member followed up with the subjects via email, text and/or phone. If follow-up emails or texts were sent, the link to the survey was included for the subjects’ convenience and to increase adherence.

The Month 9/End of Study survey was sent to subjects upon completion of the 9-month study period (270 days) or upon study withdrawal. In addition, staff completed the Study Completion/Discontinuation Form to update each subject’s study status as completed or withdrawn. The answer to the question, “Did the subject complete the study?” was critical, as it served several important functions. A ‘no’ response: 1) triggered the Month 9/End of Study survey link to be emailed to withdrawn subjects; 2) prevented remaining monthly surveys from being sent to the withdrawn subject; 3) stopped future study product shipment alerts from being sent to study staff; and 4) prevented study product reminders from being sent to subjects (discussed below). A ‘yes’ response discontinued study product reminders to subjects.

Alerts and ASIs with conditional logic were utilized throughout the REDCap project to control when or if a survey or alert was to be sent. This conditional logic used for the purposes above is considered ‘stop logic’ or a ‘kill switch’ in REDCap. An example of the Month 6 ASI conditional logic below tells REDCap to send the Month 6 survey if it has been 180 days since first dose *and* if the Study Completion/Discontinuation Form question, “Did the subject complete the study?” is yes or blank.

datediff([enrollment_arm_1][date_first_dose], ‘today’,’d’, true) = 180 and [month_9_arm_1][complete_study_eos] <> ‘0’

For access to all forms, surveys and conditional logic, see Data Availability section.

#### Control group

Toward the end of the 9-month intervention study period, survey invitations were sent using the Compose Survey Invitations feature in REDCap via the Survey Distribution Tools. No further information was required from control subjects; however, some subjects were contacted to obtain clarification on questionable data.

### Study product adherence

#### Intervention group only

Several methods were implemented to increase study product adherence including reminder emails and texts sent twice weekly; estimated self-reported vitamin D adherence on every survey; self-reported pill counts; and adherence reports for study staff review. Circular pill boxes (Pill Thing, Ellisville, MO. USA) were provided with compartments for each day of the week^20^ (Figure 7). Reminder emails included a photo of vitamin D gelcaps for picture association and automated reminder texts were sent using a text messaging service (Twilio, San Francisco, California, USA) that was integrated with REDCap. Subjects were asked to estimate the number of pills they missed on the monthly surveys, and at 3-month intervals subjects were asked to count and report the pills remaining in the study product bottles. Adherence reports in REDCap showing the self-reported estimations and pill counts were monitored; any subjects with less than 70% adherence rates were contacted to determine errors or other reasons for low adherence rates.

**Figure 7.**
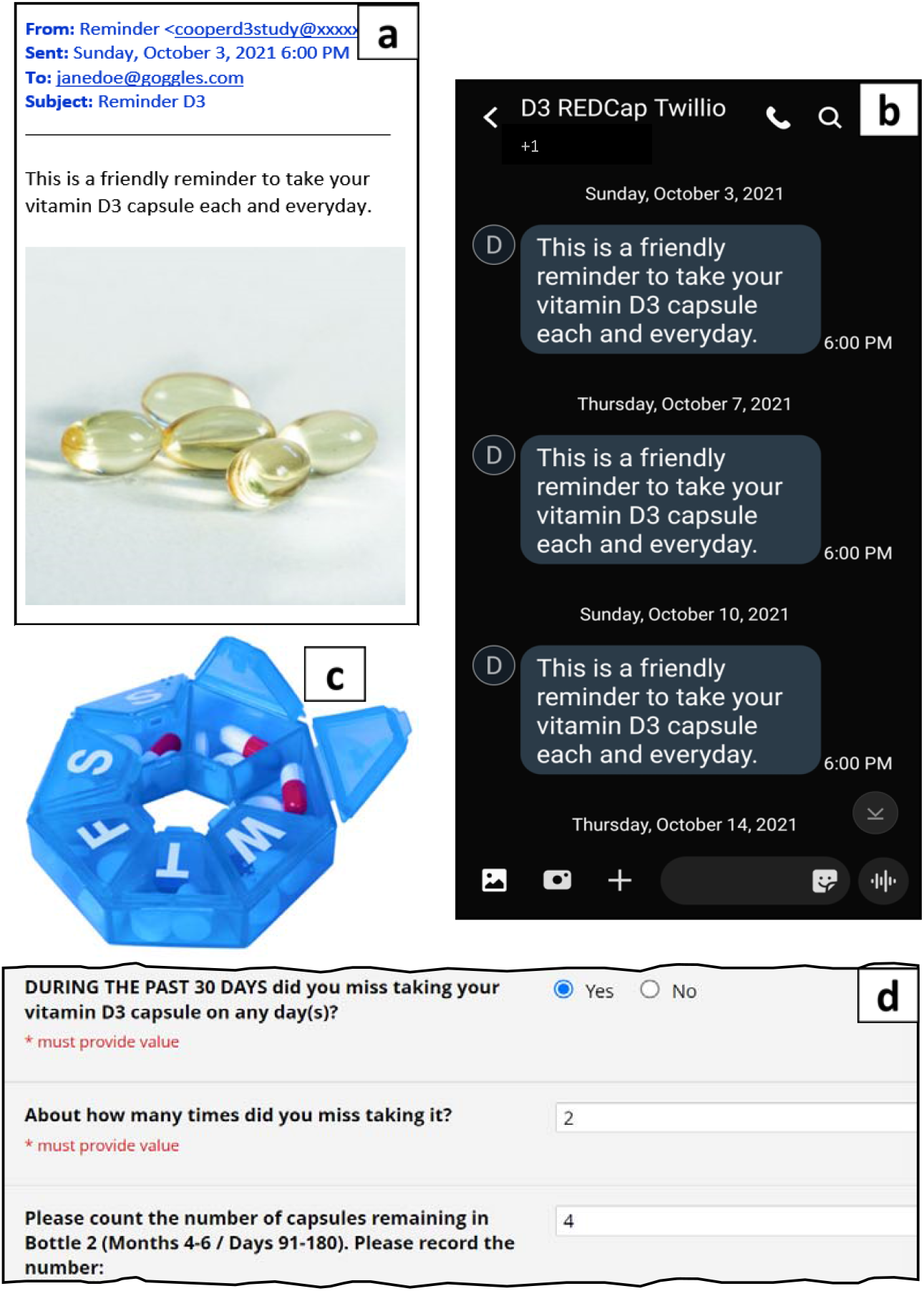
Study product adherence. To increase adherence automated email reminders (Panel a) and mobile text messages (Panel b) were sent Thursday and Sunday evenings during the subjects’ participation in the study. With the first bottle of study product, subjects were shipped 7-day pill boxes (Pill Thing, Ellisville, MO, USA) to help them remember to take the capsules (Panel c). Finally, on the monthly surveys subjects were asked if they missed taking any capsules (Panel d). In addition, subjects counted and reported remaining pills in each of the 3 bottles at months 3, 6 and 9. REDCap reports (not shown) were used to contact any subject with less than 70% adherence rate to determine errors or other reasons for low adherence rates.

REDCap’s ASIs and surveys were used unconventionally to send email and text reminders twice weekly throughout the 9-month period. Multiple surveys were created but only the ASI emails and texts were used to send the message, “This is a friendly reminder to take your vitamin D3 capsule each and every day.” The message did not include the survey link, a capability offered by REDCap ASI. Each ASI allows 1 invitation and a maximum of 5 reminders to be sent at increments of one’s choosing. Seven-day increments were chosen, thus 1 reminder could be sent weekly spanning 42 days. In order to send subjects 2 reminders (1 text and 1 email) twice weekly, a total of 28 surveys were required to span the 270-day study period. An example of the ASI conditional logic for one of the many reminders is below.

[enrollment_arm_1][date_first_dose] <> ““ and [month_9_arm_1][complete_study_eos] = ““ and datediff([enrollment_arm_1][date_first_dose], ‘today’,’d’, true) > 125 and datediff([enrollment_arm_1][date_first_dose], ‘today’,’d’, true) < 168

### Safety and adverse event monitoring

#### Intervention group only

To monitor subject safety, monthly surveys were designed to screen for potential vitamin D toxicity symptoms such as hypercalcemia and nephrolithiasis. Vitamin D laboratory test levels were not included as the safety profile for vitamin D3 at a 5,000 IU dose has been shown to be safe.^21-25^ The surveys also included questions regarding new or unusual health changes, new medications, hospitalizations and pregnancy. If any of the survey safety questions were answered “yes”, email alerts with links to the survey were triggered to the study team which enabled prompt follow up with subjects and adverse event (AE) reporting (Figure 8). If indicated, a clinic visit was offered, or laboratory tests were ordered to rule out hypercalcemia or nephrolithiasis. Documentation of physician review and assessment of the AEs was direct data entered into the ‘Physician Reviewing/Assessing AE’ field in each AE form. Documentation of subject communications were also direct data entered into the ‘Additional Comments’ field in each AE form; this field served as a charting tool. In addition, AE reports were produced weekly using REDCap’s reporting module and were distributed by email to team members to track adverse events for follow up.

**Figure 8.**
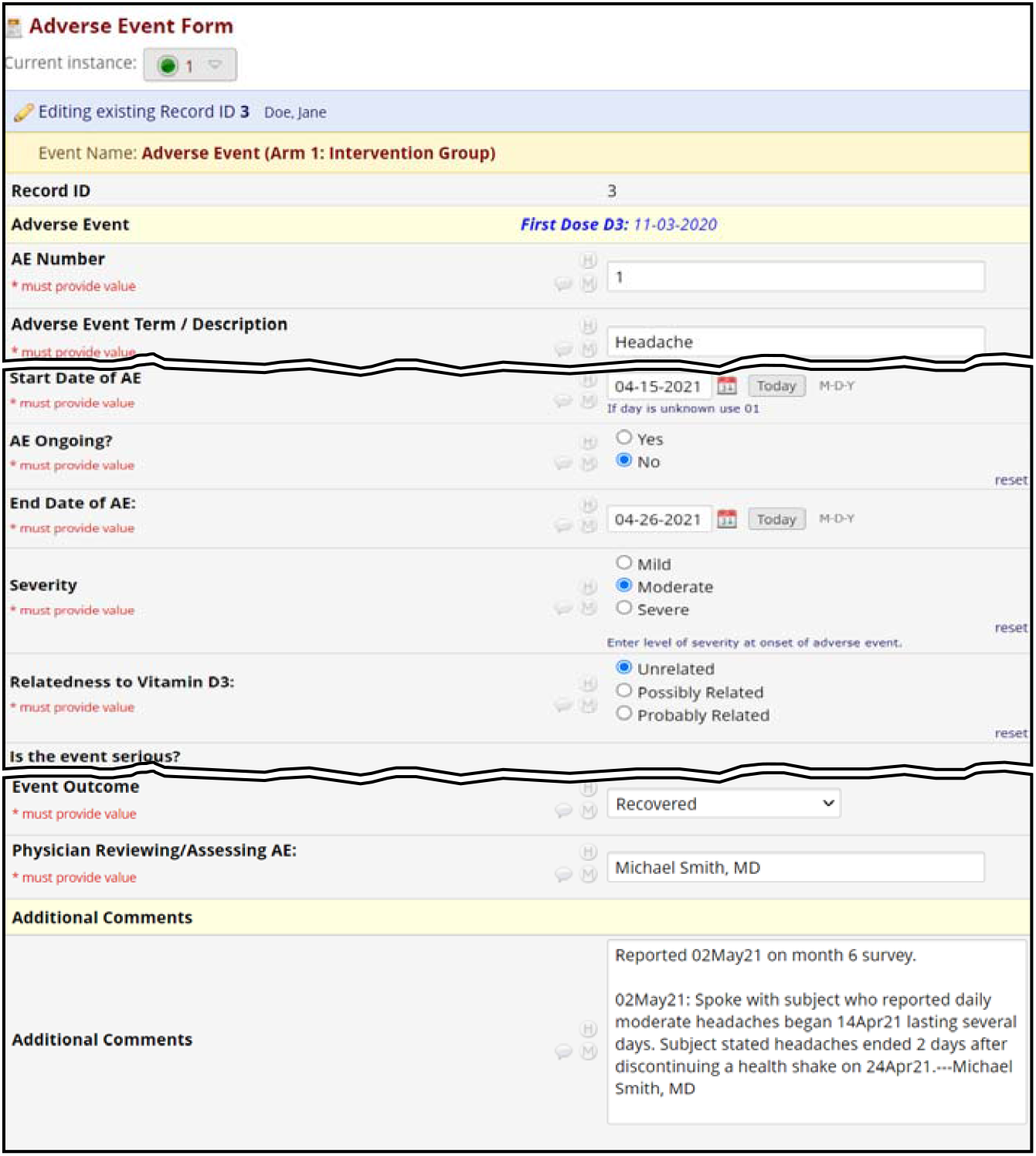
Safety monitoring / adverse events. This is a typical adverse event (AE) form that collects event term, start/end dates, severity, relatedness, seriousness and event outcome. Direct data entry was employed in lieu of paper source documentation. The “Additional Comments” field was used to document communications with the subjects. The “Physician Reviewing / Assessing AE” field was included to document physician review, relatedness assessment and adjudication. Prior to physician adjudication, this field was utilized for study team tracking to indicate which study team member was following up on a particular AE; once the AE was ready for physician review and relatedness assessment, “Complete” was entered to signal the physician to review, assess relatedness of, and adjudicate the adverse event. A weekly AE report derived from REDCap was shared with team members to track progress in AE monitoring.

A subject summary form (Figure 9) was created which pulled (or piped) pertinent subject data from other forms/surveys to provide study staff with essential information such as phone number, date of first dose, study status, medical history, survey data and adverse events with all additional comments. Staff utilized this time-saving summary prior to and during follow up phone calls with subjects. The subject summary tool was also used for quick miscellaneous reference as needed.

**Figure 9.**
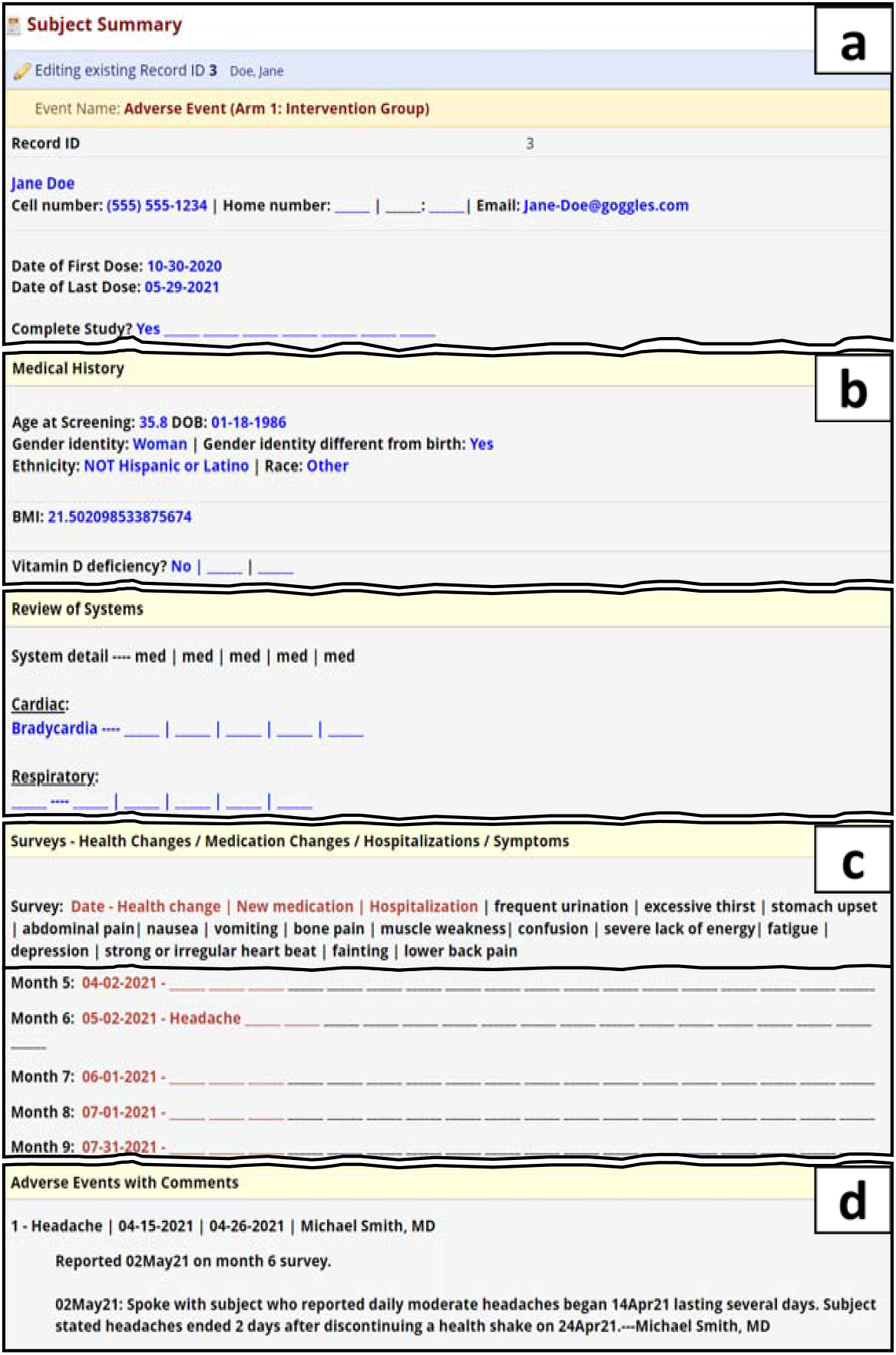
Subject summary. Utilizing the piping capability in REDCap, subject data was “piped” into this form from other surveys and forms (e.g., PHI, Demography and Medical Hx, monthly surveys, Adverse Events) to provide a summary of subject information. The summary was primarily used for reference when following up with subjects on adverse events. Panel a: The summary form streamlined communication logistics by including the subject’s name, email and phone number, along with date of first dose, last dose and subject status. Panel b: Demographics, BMI and subject-reported medical history was shown to provide background medical information. Panel c: Concise descriptors (e.g., frequent urination) for each survey question acts as a header for the piped survey responses below the header (i.e., survey name, date of survey and subject entries, if any). Panel d: Each adverse event term is listed with start/end date and additional comments.

### System usability

The System Usability Scale (SUS) is a well-recognized and standardized tool to assess perceived usability of systems.^26^ The SUS survey was sent as a REDCap survey well into the maintenance period of the study and after 57 intervention subjects completed the study. The survey was sent to 260 enrolled intervention subjects (299 less 39 subjects who had withdrawn consent). In addition, a separate SUS survey was sent to 9 of the 10 study team members to score usability from their perspectives with a focus on system usability during the recruitment, screening and informed consent process. The SUS subject survey is available in the shared XML file - see Data Availability section.

## RESULTS

### Enrollment

299 intervention subjects were enrolled in the study over a 3-month period with minimal staff consisting of 10 part-time team members. 578 control subjects voluntarily completed the control survey.

### Study product shipping

Only 2 study team members were responsible for study product shipment. Very few emails outside of REDCap alert notifications were needed when expected shipping issues arose. Out of 829 shipments, only 17 required a second bottle to be shipped. Most reasons for second shipments were carrier delays or lost shipment; other reasons included address changes and damaged product.

### Data completeness

The average adherence rate for completing monthly surveys was 98.5%; the high rate is attributed to automated reminders and manual follow up. A dedicated staff member manually followed up with emails, texts and/or phone calls if subjects did not complete surveys after the initial automated survey invitation and 3 daily reminder emails were sent. Throughout the 9-month study period, a total of 458 texts and 616 emails were sent and 208 phone calls were made to boost survey adherence.

### Study product adherence

The average self-reported vitamin D adherence rate was 87%.

### Safety monitoring

The automated alerts, direct data entry of subject communications, the subject summary tool and weekly AE reports were the tools that improved efficiency and safety data accuracy and allowed 3 team members to effectively monitor and follow 388 adverse events over the course of the study. Three in-person clinic visits occurred and 22 laboratory tests (vitamin D, calcium, phosphorus, magnesium levels) were ordered as a result of adverse events follow up.

### System usability

SUS survey data assessed system usability from both the subject and study staff perspectives. Out of 260 subject surveys sent, 196 subjects responded with an average score of 93.8%. All 9 staff members who were sent the SUS survey responded with an average score of 90% when considering the usability during recruitment, screening and electronic consent.

## DISCUSSION

### Principal findings

The novel internet methods developed to conduct a randomized interventional controlled trial allow for efficient subject recruitment with excellent study data completeness. Our trial was designed to be conducted remotely utilizing the REDCap data capture platform and vCita electronic scheduling application. We heavily relied on electronic methods to efficiently and cost-effectively conduct a clinical trial while avoiding being a burden on the community or potentially contributing to community spread of COVID-19. The remote methods significantly reduced the costs of conducting a clinical trial.^2,8^ The methods enabled enrollment of 299 subjects over 3 months with only 10 part-time study staff members. In addition, facility space to interview subjects was unnecessary as all screening and consenting was performed remotely. Storage space for all the typical paperwork was greatly reduced. Additional savings are noted in that staff and subjects did not need to travel to and from a facility. Electronic consenting saved on staff that would have been needed to review and archive paper consents. Time was also saved with e-consenting because the auto date- and time-stamping feature in REDCap eliminated errors that are common with traditional paper informed consents.^27^

Remote and automated electronic methods were utilized by a small group of study staff to successfully recruit, consent and enroll 299 intervention subjects, and collect 578 control surveys. By applying complex conditional logic in REDCap, efficient automation of surveys, alerts, product shipment notifications and study product reminders was attained. In addition to receiving high system usability scores (SUS), the methods used produced excellent adherence rates in both subject study product compliance and survey completion. The electronic tools developed in REDCap for data collection and safety monitoring and overall study maintenance reduced the extensive time, costs and staff typically necessary to conduct a clinical trial.

Vitamins and supplements are rarely studied partly due to the cost needed to design and conduct a randomized clinical study. These methods can improve subject recruitment, subject monitoring, data collection in trials outside corporate funded trials.

### Limitations/Future directions

We encountered challenges with recruitment that resulted in low enrollment numbers. We surmised that many potential participants were lost due to the incompatibility between outdated browser software and the 3^rd^ party scheduling application. Although our institutional IT department confirmed all browser software had been updated with the latest version, we found that individual subjects were still using the outdated browser on their desktops which caused problems with electronically scheduling screening appointments. Other lessons learned are described in Supplement 2.

Many users find creating a project in REDCap daunting. REDCap is being used in innovative ways for purposes other than research among institutions.^28^ Many articles are available that share detailed information and data such as REDCap conditional logic,^13^ trial flowcharts and lessons learned,^9,12^ and practical examples such as XML data files^11^ which will give a novice user a complete REDCap project to use as a template to develop their own projects with ease. Access to the XML file for this project is described in the Data Availability section.

RedCAP is an exceptional data collection tool that is only available to nonprofit institutions. Research equity would be more impactful were it made available to private practices.

### Conclusions

The remote and automated methods developed for this randomized clinical trial yielded efficient subject recruitment with excellent study data completeness and a significant reduction in operational costs without sacrificing safety or quality. These shared methods offer researchers the means to conduct their own clinical trials efficiently and cost-effectively.

## Data Availability

The datasets generated during and/or analyzed during the current study are available from the corresponding author following reasonable request.

## Abbreviations

REDCap: Research Electronic Data Capture
COVID-19: coronavirus disease 2019
ASI: Automated Survey Invitations
AE: Adverse Event
SUS: System Usability Scores
PHI: Protected Health Information

## ACKNOWLEDGEMENTS

We extend gratitude to Susan J Lamon PhD for her advice on reminders to promote vitamin D3 adherence and to all the healthcare workers who were willing to participate in this study.

## DATA AVAILABILITY

An XML file containing our REDCap project metadata is available for upload into the reader’s own REDCap application (Supplement 3). Subject data is not included in this XML file. The metadata includes user roles, record status dashboards, reports, alerts and notifications, surveys and survey settings, and automated survey invitations. We share this file to help other researchers observe how REDCap was used in our trial, learn from it, or use it as a starting point in creating their own study. The XML file can be uploaded simply by following these steps: 1) download the XML file; 2) create a new project in REDCap; 3) in the ‘Project creation option’ select ‘Upload a REDCap project XML file’; and 4) select ‘Choose File’ and locate and open the XML file that was downloaded in step 1.

## Contributors

TC wrote the initial draft of this article. NVH, BB, PJL and LVM were responsible for critical review of the article and for important intellectual content. NVH and PJL wrote the protocol and were responsible for the design of the methods. Conceptualization of the study included NVH, TLB, PJL, SR, BB, LVM and MKC. NVH, PJL, TC, BB, LVM and HG were responsible for project administration. NVH, TLB, PJL, TC, SR, BB, KQN, HG, LMV, AT, DT and MAM were responsible for data collection, and review and editing of the article. KH and MKC reviewed and commented on the article. TC was responsible for the creation of the database for data collection. TC, HG, DT and MAM were responsible for data management. NVH and TC calculated results for analyses. NVH and KH completed the data analyses. KH validated results. TC and BB were responsible for the management of software used in subject recruitment and data collection. All authors had access to the final study results and were responsible for the final approval of the manuscript.

## SUPPLEMENTAL INFORMATION

**Supplement 1** Electronic informed consent signing

**Supplement 2** Lessons Learned

**Supplement 3** XML File

