## Supplement 1 for "Remote and semi-automated methods to conduct a randomized clinical trial" (from another version of this preprint: version 1, 2022-11-11)

### Supplement 1 - eSigning Informed Consent

The following are steps taken by the subject and investigator to electronically sign the informed consent remotely whilst utilizing the informed consent framework in REDCap. Our automated survey invitation (ASI) conditional logic is set to email the subject a link to the informed consent [survey] after the investigator answers ‘yes’ to the eligibility question on the Screening and Eligibility form. The subject clicks on the link to the informed consent to open the informed consent. At the same time, the investigator opens the subject’s informed consent from the Record Status Dashboard or Add/Edit Records page in REDCap (i.e., both the subject and the investigator are viewing the informed consent at the same time). After the informed consent is reviewed with the subject and all questions are answered, both parties sign the consent by following the steps below in order.

#### Subject:

Reads and completes Voluntary Participation section and clicks ‘yes’ or ‘no.’

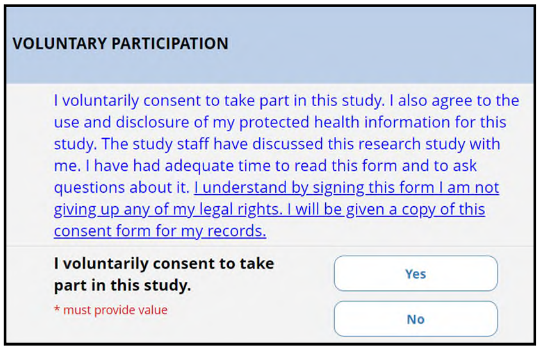


Completes Subject Signature section and enters name and email; signs by clicking the ‘Add signature’ link; and enters date and time by selecting the ‘Now’ button.

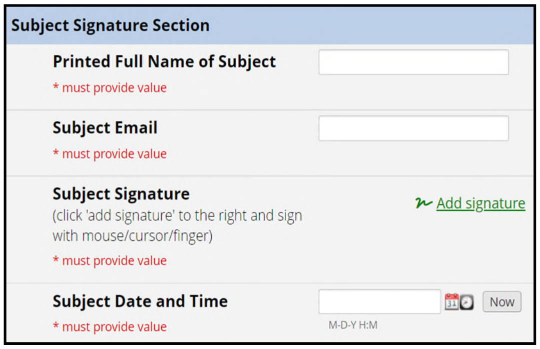


Scrolls to bottom and clicks ‘Save & Return Later.’ This allows the investigator to sign his/her portion of the consent.

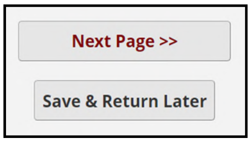

Closes small ‘Return Code needed to return’ window, but stays on browser page.

(If subject inadvertently exits page he/she can reopen the survey from the email and access by entering the return code. The investigator can provide the return code.)

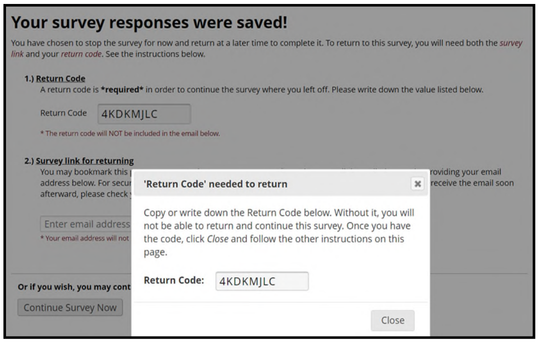


### Investigator:

Opens subject informed consent from the Record Status Dashboard or Add/Edit Records page in REDCap.

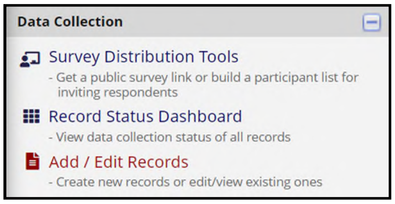


Refreshes consent page (browser page) which enables investigator to confirm subject signature section was completed.

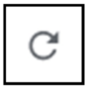

Clicks on ‘Edit response’ to make the survey editable.

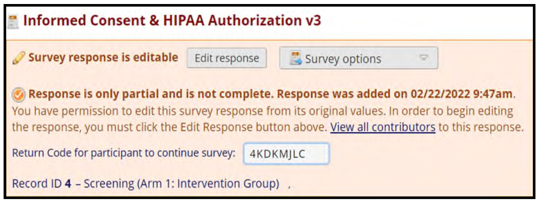


Reads and ticks ‘Investigator statement’; enters name; signs by clicking the ‘Add signature’ link; and enters date and time by selecting the ‘Now’ button.

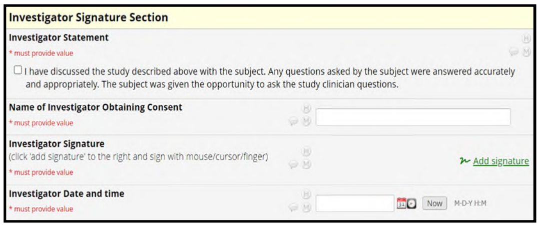


Selects ‘Save and Stay’

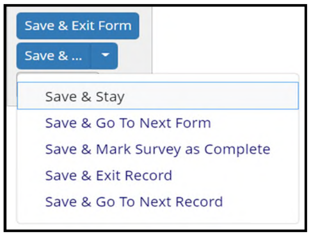

### Subject:

Clicks on ‘Continue Survey Now’

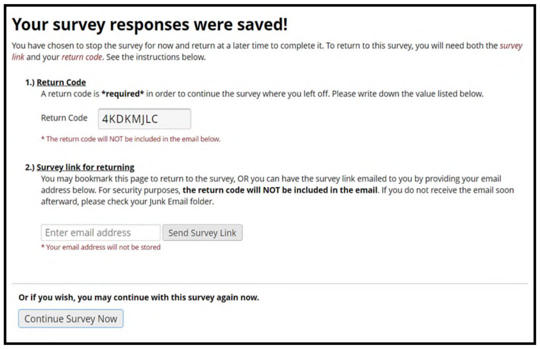


Scrolls to bottom of consent and clicks ‘Next Page’

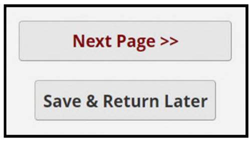


Reviews inline consent and scrolls down to confirm fully signed.

Downloads or prints consent for their records.

Certifies the document by ticking the yellow certification statement.

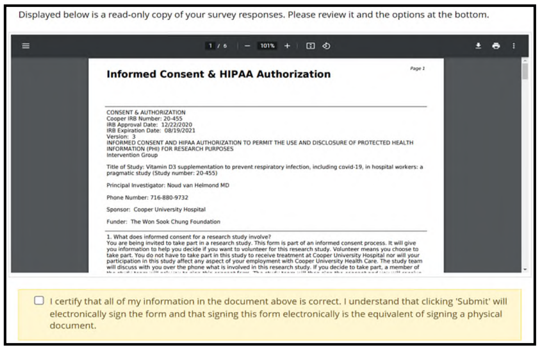


Sees certification statement changes to green once ticked.

Clicks ‘Submit.’

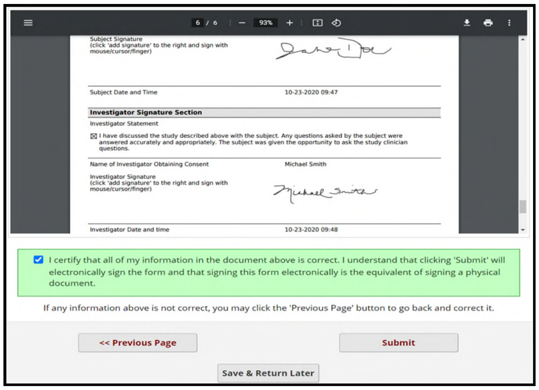


Ticks ‘Close survey’ or may download consent from this page if issues were encountered in the previous screen.

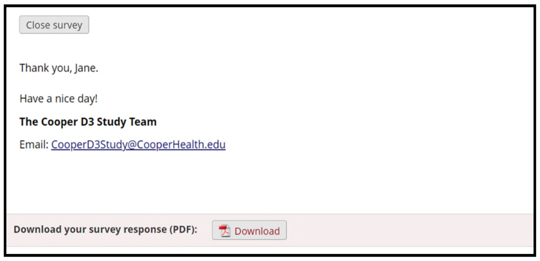

### Investigator:

Refreshes consent page (browser page).

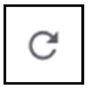

Reviews completed consent and confirms no information (names, dates/times, signatures) was changed.

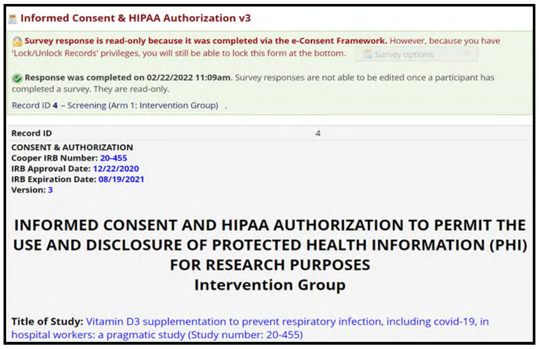

**Informed Consent & HIPAA Authorization v3**

**Survey response is read-only because it was completed via the e-Consent Framework.** However, because you have 'Lock/Unlock Records' privileges, you will still be able to lock this form at the bottom. Survey options

**Response was completed on 02/22/2022 11:09am.** Survey responses are not able to be edited once a participant has completed a survey. They are read-only.

Record ID **4** – Screening (Arm 1: Intervention Group) .

| | |
|---|---|
| **Record ID** | 4 |

**CONSENT & AUTHORIZATION**
**Cooper IRB Number:** 20-455
**IRB Approval Date:** 12/22/2020
**IRB Expiration Date:** 08/19/2021
**Version:** 3

**INFORMED CONSENT AND HIPAA AUTHORIZATION TO PERMIT THE USE AND DISCLOSURE OF PROTECTED HEALTH INFORMATION (PHI) FOR RESEARCH PURPOSES**
**Intervention Group**

**Title of Study:** Vitamin D3 supplementation to prevent respiratory infection, including covid-19, in hospital workers: a pragmatic study (Study number: 20-455)
