## Supplement 2 for "Remote and semi-automated methods to conduct a randomized clinical trial" (from another version of this preprint: version 1, 2022-11-11)

### Supplement 2 - Lessons Learned

**Data Dictionary:** We experienced difficulty when trying to upload an updated data dictionary or automated survey invitation file. We used MS Excel to make updates to these comma delimited .csv data sets, however, when we attempted to upload, REDCap would yield errors. After lengthy research the culprit was found to be the smart quotes in Excel are not readable by REDCap. The very easy fix to this was to make the changes in Excel, but before uploading, open the file as a Google sheet and download it as a .csv file. Uploading a .csv file produced by Google Sheets did not trigger errors. Another option not tried would have been to disable the smart quotes in Excel.

**Alerts** would have been more efficient when setting up reminders for the project, however at the time the team was working with an outdated version of REDCap that did not offer alerts.

**Number of events** in longitudinal study – We made a separate event for each of the 10 surveys. In hindsight, the surveys could have been placed under 1 event and bottle shipment surveys and forms under another. This would have made data reports for analysis less cumbersome and navigation easier. The figure on the left is our events schedule; the figure on the right is more desirable in terms of navigation between forms/surveys. In addition the event schedule on the right would have made conditional logic writing much easier.

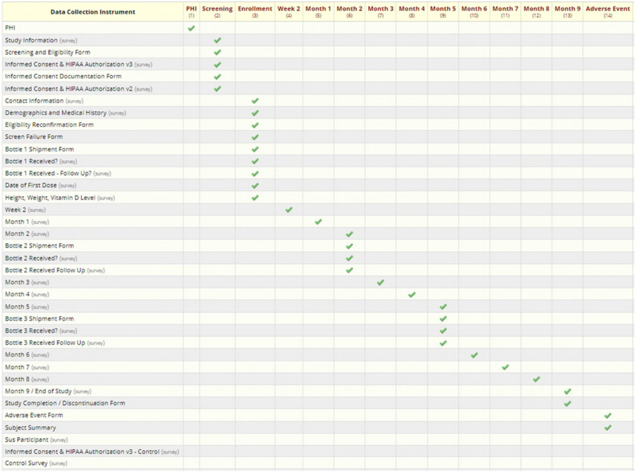

| Data Collection Instrument | PHI (1) | Screening (2) | Enrollment (3) | Week 2 (4) | Month 1 (5) | Month 2 (6) | Month 3 (7) | Month 4 (8) | Month 5 (9) | Month 6 (10) | Month 7 (11) | Month 8 (12) | Month 9 (13) | Adverse Event (14) |
|---|---|---|---|---|---|---|---|---|---|---|---|---|---|---|
| PHI | ✓ | | | | | | | | | | | | | |
| Study Information (survey) | | ✓ | | | | | | | | | | | | |
| Screening and Eligibility Form | | ✓ | | | | | | | | | | | | |
| Informed Consent & HIPAA Authorization v3 (survey) | | ✓ | | | | | | | | | | | | |
| Informed Consent Documentation Form | | ✓ | | | | | | | | | | | | |
| Informed Consent & HIPAA Authorization v2 (survey) | | ✓ | | | | | | | | | | | | |
| Contact Information (survey) | | | ✓ | | | | | | | | | | | |
| Demographics and Medical History (survey) | | | ✓ | | | | | | | | | | | |
| Eligibility Reconfirmation Form | | | ✓ | | | | | | | | | | | |
| Screen Failure Form | | | ✓ | | | | | | | | | | | |
| Bottle 1 Shipment Form | | | ✓ | | | | | | | | | | | |
| Bottle 1 Received? (survey) | | | ✓ | | | | | | | | | | | |
| Bottle 1 Received - Follow Up? (survey) | | | ✓ | | | | | | | | | | | |
| Date of First Dose (survey) | | | ✓ | | | | | | | | | | | |
| Height, Weight, Vitamin D Level (survey) | | | ✓ | | | | | | | | | | | |
| Week 2 (survey) | | | | ✓ | | | | | | | | | | |
| Month 1 (survey) | | | | | ✓ | | | | | | | | | |
| Month 2 (survey) | | | | | | ✓ | | | | | | | | |
| Bottle 2 Shipment Form | | | | | | ✓ | | | | | | | | |
| Bottle 2 Received? (survey) | | | | | | ✓ | | | | | | | | |
| Bottle 2 Received Follow Up (survey) | | | | | | ✓ | | | | | | | | |
| Month 3 (survey) | | | | | | | ✓ | | | | | | | |
| Month 4 (survey) | | | | | | | | ✓ | | | | | | |
| Month 5 (survey) | | | | | | | | | ✓ | | | | | |
| Bottle 3 Shipment Form | | | | | | | | | ✓ | | | | | |
| Bottle 3 Received? (survey) | | | | | | | | | ✓ | | | | | |
| Bottle 3 Received Follow Up (survey) | | | | | | | | | ✓ | | | | | |
| Month 6 (survey) | | | | | | | | | | ✓ | | | | |
| Month 7 (survey) | | | | | | | | | | | ✓ | | | |
| Month 8 (survey) | | | | | | | | | | | | ✓ | | |
| Month 9 / End of Study (survey) | | | | | | | | | | | | | ✓ | |
| Study Completion / Discontinuation Form | | | | | | | | | | | | | ✓ | |
| Adverse Event Form | | | | | | | | | | | | | | ✓ |
| Subject Summary | | | | | | | | | | | | | | ✓ |
| Sus Participant (survey) | | | | | | | | | | | | | | |
| Informed Consent & HIPAA Authorization v3 - Control (survey) | | | | | | | | | | | | | | |
| Control Survey (survey) | | | | | | | | | | | | | | |

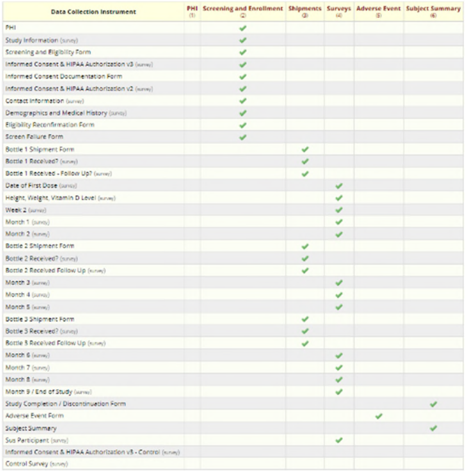

| Data Collection Instrument | PHI (1) | Screening and Enrollment (2) | Shipments (3) | Surveys (4) | Adverse Event (5) | Subject Summary (6) |
|---|---|---|---|---|---|---|
| PHI | | ✓ | | | | |
| Study Information (survey) | | ✓ | | | | |
| Screening and Eligibility Form | | ✓ | | | | |
| Informed Consent & HIPAA Authorization v3 (survey) | | ✓ | | | | |
| Informed Consent Documentation Form | | ✓ | | | | |
| Informed Consent & HIPAA Authorization v2 (survey) | | ✓ | | | | |
| Contact Information (survey) | | ✓ | | | | |
| Demographics and Medical History (survey) | | ✓ | | | | |
| Eligibility Reconfirmation Form | | ✓ | | | | |
| Screen Failure Form | | ✓ | | | | |
| Bottle 1 Shipment Form | | | ✓ | | | |
| Bottle 1 Received? (survey) | | | ✓ | | | |
| Bottle 1 Received - Follow Up? (survey) | | | ✓ | | | |
| Date of First Dose (survey) | | | | ✓ | | |
| Height, Weight, Vitamin D Level (survey) | | | | ✓ | | |
| Week 2 (survey) | | | | ✓ | | |
| Month 1 (survey) | | | | ✓ | | |
| Month 2 (survey) | | | | ✓ | | |
| Bottle 2 Shipment Form | | | ✓ | | | |
| Bottle 2 Received? (survey) | | | ✓ | | | |
| Bottle 2 Received Follow Up (survey) | | | ✓ | | | |
| Month 3 (survey) | | | | ✓ | | |
| Month 4 (survey) | | | | ✓ | | |
| Month 5 (survey) | | | | ✓ | | |
| Bottle 3 Shipment Form | | | ✓ | | | |
| Bottle 3 Received? (survey) | | | ✓ | | | |
| Bottle 3 Received Follow Up (survey) | | | ✓ | | | |
| Month 6 (survey) | | | | ✓ | | |
| Month 7 (survey) | | | | ✓ | | |
| Month 8 (survey) | | | | ✓ | | |
| Month 9 / End of Study (survey) | | | | ✓ | | |
| Study Completion / Discontinuation Form | | | | | | ✓ |
| Adverse Event Form | | | | | ✓ | |
| Subject Summary | | | | | | ✓ |
| Sus Participant (survey) | | | | ✓ | | |
| Informed Consent & HIPAA Authorization v3 - Control (survey) | | | | | | |
| Control Survey (survey) | | | | | | |

**Navigation:** To quickly jump from one subject survey or form to another one can simply locate the subject number in the URL for a particular subject survey or form and change the subject number to the desired subject number. This eliminates the multiple steps and time needed to access a subject record through the Record Status Dashboard or the Add / Edit Records in the Data Collection menu. For example, the following URL was saved as a browser favorite titled MedHx. One only had to overwrite the XXXX with the subject record number to display that subject's Medical History survey.

https://redcap.rowan.edu/redcap_v10.8.2/DataEntry/index.php?pid=208&id=20-XXXX&event_id=668&page=demographics_and_medical_history

**AE Comment Field:** We utilized an Adverse Event (AE) comment field to document communications with subjects using direct data entry. In hindsight, creating a separate form acting as progress notes may have been more efficient.

**Reminders:** Explain issue with reminders logic that prevented reminders from being sent. (From Bangdiwala 'Technical procedures and REDCap tools for internet-based clinical trials' article: When the "ensure logic is true before sending automated invitation" option is selected, "REDCap will re-evaluate the logic against the record's data values whenever the record values are changed AFTER the invitation has been scheduled but BEFORE it has been sent to the respondent.")

**Shipment tracking link** was provided to subjects if they answered 'no' on the survey. In hindsight, we should have made the link available on the survey without the need to use branching logic.
